# Characteristics and antiviral treatment eligibility of newly diagnosed hepatitis B patients at a teaching hospital in Ghana: Implications for prevention and management

**DOI:** 10.1101/2024.03.28.24305058

**Authors:** Joseph Daniels, Yvonne A. Nartey, Francis Djankpa, Jacques Simpore, Dorcas Obiri-Yeboah

**Author notes:** Corresponding author (JD).

## Abstract

Hepatitis B virus (HBV) infection poses a considerable public health challenge in limited-resource settings especially in the sub-Saharan African region. Even though HBV infection is incurable, timely treatment is effective in preventing disease progression to liver cirrhosis or hepatocellular carcinoma. However, not all infected patients require treatment. The aim of this study was to determine the clinical, immunological, and virological profiles of newly diagnosed adult HBV patients at a tertiary healthcare center in Ghana and to determine the antiviral treatment eligibility rate based on current guidelines of the World Health Organization (WHO). A hospital-based cross-sectional study involving total sampling of 220 treatment naïve HBV surface antigen positive clients was carried out. A structured questionnaire was used to collect data and detailed clinical and laboratory assessment (serological, biochemical and virological) was carried out. Data were entered and analyzed with STATA version 16. The median age at diagnosis was 34 years (IQR 26.0 – 41.5) with a male to female ratio of 1:1.5. A total of 138 participants (62.7%) were diagnosed with HBV infection following voluntary testing. There was a median delay of 8.5 months (IQR 3.0 – 22.5) between initial diagnosis and patients’ presentation for medical care. In all, 24 patients (10.9%) had abnormal clinical examination findings, 172 patients (78.2%) had HBV DNA levels ≤ 2000 IU/ml while 8 (3.6%) were seropositive for HBeAg. A small proportion of patients had concomitant human immunodeficiency virus (2.7%) and hepatitis C virus (1.4%) infections. Treatment eligibility rate was very low among newly diagnosed HBV infected patients seeking medical care (n=14, 6.4%) following the WHO guidelines for treatment eligibility. Thus, increasing screening rate among the general population, early linkage to clinical care of screen positives and vaccination of screen negatives will help reduce HBV related clinical conditions in resource limited countries.

## Introduction

Estimated global prevalence rates in 2022, based on data from 170 countries suggest that about 257.5 million individuals worldwide have confirmed hepatitis B virus (HBV) infection [1]. About 0.5% of infected people yearly seroconvert spontaneously from being hepatitis B surface antigen (HBsAg) positive to becoming hepatitis B surface antibody (HBsAb) positive [2]. The overall global lifetime risk of HBsAg seropositivity is estimated to be more than 60% [3]. The World Health Organization (WHO) estimates the occurrence of 1.5 million new infections around the globe every year. HBV infects the liver and is known to be a major global cause of both acute and chronic hepatic diseases [4]. HBV infection is also reported to be the cause for about 50 - 80% of all diagnosed cases of hepatocellular carcinomas (HCC) globally [5]. Additionally, it is estimated to cause between 500,000 and 1.2 million deaths worldwide annually through chronic hepatitis, and associated complications such as liver cirrhosis and HCC [6].

HBV infection poses a considerable public health challenge in the developing world especially African countries [7]. In Africa unlike other regions of the world, HCC and liver cirrhosis develop at an earlier age among patients with HBV infection, and are generally associated with poorer prognosis [8]. The highest prevalence of HBsAg seropositivity is found in the WHO Western Pacific Region and Africa with 116 million and 81 million people respectively reported to be chronically infected [4]. In Ghana, there is high (>8%) prevalence of hepatitis B infection per the WHO classification [9]. Different experts have reported national prevalence rates of 8.48% [10], 10-15% [11] as well as 12.92% [2]. However, a recent meta-analysis of 21 studies conducted in Ghana between 2015 and 2019 reported hepatitis B infection rates of 8.36% and 14.3% among adults and adolescents respectively [12]. HBV infection requires great attention in Ghana due to its significant public health importance. HBsAg detection remains the primary tool for the screening of individuals for HBV in sub-Saharan Africa [13]. A positive test for HBsAg for more than 6 months coupled with seropositivity for hepatitis B core immunoglobulin G (HBcAb – IgG) imply disease chronicity with increased risk of developing cirrhosis, hepatic decompensation and HCC [14].

Not all patients who test positive for HBsAg require treatment. Patients’ eligibility for the commencement of antiviral therapy is dependent on key factors such as age, HBV viral load, serum Alanine Transaminase (ALT) levels, Aspartate to Platelet Ratio Index (APRI), clinical evidence of liver cirrhosis, hepatitis B envelope antigen (HBeAg) status, concomitant hepatitis C virus (HCV) or human immunodeficiency virus (HIV) coinfection and pregnancy status among females [15, 16]. It is important to accurately classify patients based on these parameters, since this directly influences clinical decisions regarding antiviral therapy. These parameters are also important in identifying patients at risk of progression to liver cirrhosis or cancer, and as such determine which individuals require immediate initiation of antiviral therapy.

With the high endemicity of chronic HBV (CHB) infection in Ghana, it is important to determine the proportion of HBV patients who require treatment. It is equally important to identify those who present for the first time with complications such as cirrhosis, hepatic decompensation and HCC, since this highlights missed opportunities for the care and management of persons living with HBV. The aim of this study was to determine the clinical, immunological, and virological profiles of newly diagnosed adult HBV infected patients at a tertiary healthcare facility in Ghana and to determine the antiviral treatment eligibility rate based on WHO guidelines.

## Methods

### Study design and setting

This was a hospital-based cross-sectional study. Participants were recruited into the study from the viral hepatitis/ sexually transmitted infections (STIs) clinic at the Cape Coast Teaching Hospital (CCTH) which is situated at the northern part of the Cape Coast metropolis in the Central Region of Ghana. CCTH is a 400-bed capacity hospital that serves as the tertiary care referral center for both private and public healthcare institutions in the central and western region of Ghana. The clinic started operating in 2009 and had registered about 3000 clients with HBV infection by 2021. CCTH provides comprehensive HBV care and runs a specialist-led gastrointestinal (GI) and hepatology clinic that manages patients with diseases of the GI and hepatobiliary tracts including those with complications related to viral hepatitis such as liver cirrhosis and hepatocellular carcinoma.

### Study Participants

Participants were recruited over a cumulative period of 11 months from December 10, 2019 to June 09, 2020 and November 03, 2020 to February 09, 2021 due to interruptions to the operations of the clinic caused by the COVID-19 pandemic. Samples were collected at the clinic once a week. There was total population sampling of all newly diagnosed clients with HBV infection who were seeking clinical care at CCTH during the sampling period. Patients included in this study were treatment naïve adult clients who were 18 years or older, with a positive HBsAg test who provided written informed consent. All patients with known HBV/HIV coinfection who were already on highly active antiretroviral therapy (HAART) were excluded from the study as well as individuals on antiviral therapy for other conditions if their medications were known to be simultaneously active against HBV infection. All patients who had been previously treated for HBV infection were also excluded from the study.

### Study size

All clients attending the clinic who met the inclusion criteria (n=231 over the recruitment period) were offered the opportunity to be part of the study. However, 11 (4.8%) eligible clients declined to provide informed consent to participate in the study for personal reasons. In all, 220 patients were recruited into the study.

### Ethical considerations

The study was approved by the institutional review board of CCTH (Reference number: CCTHERC/EC/2019/084). All patients’ information including laboratory test results were kept private and confidential. The data of the participants were anonymized prior to analysis. Throughout this study, there was strict adherence to the prescribed standards of acceptable scientific and ethical behavior. All relevant COVID-19 protocols were observed during interactions with patients and data collection. All the study participants provided written informed consent prior to their recruitment into the study.

### Data sources

Data regarding socio-demographic characteristics, past medical and family history as well as other relevant factors linked to HBV infection were collected through a structured questionnaire which comprised 32 closed-ended questions. Relevant data were also abstracted from patients’ hospital records with their consent. The questionnaire was pretested in a similar out-patient setting to determine its validity and reliability to ascertain its appropriateness for this study. Twenty-two (22) clients were selected for pretesting, representing 10% of the actual sample size. Tau-equivalent reliability, also known as Cronbach’s Alpha (α) was computed to determine the reliability coefficient. The Alpha value obtained was 0.72 (number of items = 32), therefore this research instrument was deemed reliable and acceptable for collecting useful data for the study.

### Laboratory Procedures / Analysis

Single-use commercially available First Response® HBsAg rapid test kits (Premier Medical Corporation Private Limited, India) were used to qualitatively detect HBsAg in the plasma specimen of patients [17]. Using the COBAS® AmpliPrep / COBAS® TaqMan® HBV Test, version 2.0 (Roche, Switzerland) (CAP-CTM) and in accordance with the manufacturer’s instructions, the HBV viral load was measured from 400µl of patients’ blood [18]. The Biopanda HBV Combo Rapid Test (RAPG-HBV-001) (Biopanda, United Kingdom) was utilized to qualitatively detect HBsAg, HBsAb, HBeAg, HBeAb, and HBcAb in patients’ blood samples. The ChemWell® 2910 Automated EIA and Chemistry Analyzer (Awareness Technologies, USA) was used to measure serum electrolytes, blood urea nitrogen (BUN) and liver enzymes including aspartate and alanine transaminases, strictly following the manufacturer’s protocol. Complete blood counts (CBCs) were determined using the Mindray BC-2800 Auto Hematology Analyzer (China) [19]. All the patients were also tested for HIV infection using the First Response^®^ HIV 1-2.O test (Premier Medical Corporation Private Limited, India), OraQuick**^®^** HIV test (OraSure Technologies, USA) and SD Bioline™ HIV-1/2 antibody test kits (Abbott, USA) based on the guidelines for HIV screening and diagnosis among adults, pregnant women and adolescents in healthcare settings [20]. Single use commercially available HCV rapid test kits (Advanced Diagnostic Kit for immunoglobulin to HCV, China) were used to qualitatively detect the presence of anti-HCV immunoglobulins in patients’ samples with sensitivity of 99.0% and specificity of 99.8% [21]. APRI calculation was based on hematological and biochemical tests conducted on patients’ samples. APRI was computed with the formula (AST/40/ platelet count (10^9^/L) x 100) [22].

### Statistical analysis

Data were anonymized, coded, statistically cleaned, and analyzed using STATA statistical software package version 16 for Microsoft windows (Stata Corp). A combination of descriptive and inferential statistics was performed. Socio-demographic and other characteristics of participants were presented as frequencies with percentages. Histogram and Shapiro-Wilk’s test were used to assess normality of the data. Means and standard deviations were used for normally distributed data while median and interquartile range were used for skewed data. For this study, *p* ≤ 0.05 was considered statistically significant at a 95% confidence interval.

## Results

### Socio-demographic and behavioral characteristics

In total, 220 individuals with confirmed hepatitis B viral infection participated in this study. A total of 131 (59.6%) of the participants were female whereas 109 (40.4%) were male. There were more females than males in all age categories (Fig 1).

**Fig 1.**
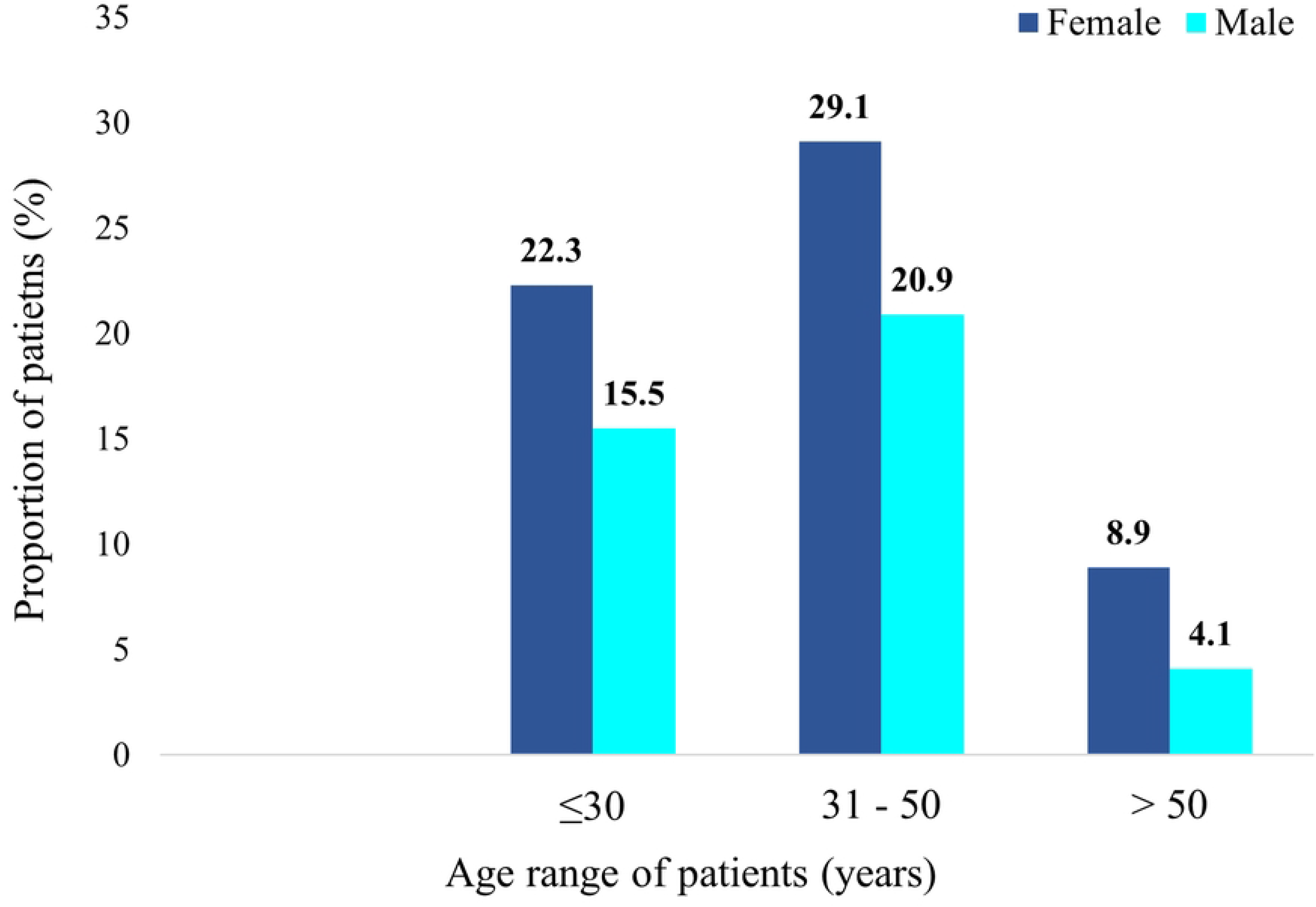
Percentage distribution of patients by age and gender.

The median age was 34 years (IQR 26.0 – 41.5). One hundred and eight (49.0%) were single whilst the remaining (n=112, 51.0%) were either married, divorced, or widowed. Majority (n=143, 65.0%) were sexually active. The median interval between initial diagnosis and presentation at the clinic was 8.5 months (IQR 3.0 – 22.5), with about a third of the patients (61.8%) attending their first visit to the clinic more than a year after initial diagnosis (Table 1).

**Table 1.** Socio-demographic and behavioral characteristics (n=220)

| **Variables** | **Categories** | **Number (n)/**<br>**Median** | **Percentage (%)/**<br>**IQR** |
| --- | --- | --- | --- |
| **Age (years)** | Median | 34 | 26.0 – 41.5 |
|  | ≤ 30 | 83 | 37.7 |
|  | 31 – 50 | 110 | 50.0 |
|  | > 50 | 27 | 12.3 |
| **Marital status** | Single | 108 | 49.0 |
|  | Married | 100 | 45.5 |
|  | Divorced/<br>Widowed | 12 | 5.5 |
| **Sexual activity** | Active | 143 | 65.0 |
|  | Inactive | 77 | 35.0 |
| **Pregnancy status of**<br>**females (n = 131)** | Pregnant | 9 | 6.9 |
|  | Not-pregnant | 122 | 93.1 |
| **Time since initial**<br>**diagnosis (years)** | Median | 8.5 | 3.0 – 22.5 |
|  | ≤ 1 | 136 | 61.8 |
|  | 1 – 5 | 65 | 29.6 |
|  | > 5 | 19 | 8.6 |
| **History of alcohol**<br>**intake** | Yes | 30 | 13.6 |
|  | No | 190 | 86.4 |

| Variables | Categories | Number (n)/<br>Median | Percentage (%)/<br>IQR |
| --- | --- | --- | --- |
| Smoking habit | Smokers | 6 | 2.7 |
|  | Non-smokers | 214 | 97.3 |

IQR: interquartile range

Most of the participants (n = 138, 62.7%) were initially diagnosed with HBV infection due to voluntary testing whereas 19 females (8.6%) were diagnosed as part of routine ANC screening. A few (n = 8, 3.6%) were diagnosed through screening after the diagnosis of close associates (sexual partners & family members) whereas the remainder (n=55, 25.0%) were diagnosed after testing pursuant to the recommendation of a health-worker (Fig 2).

**Fig 2.**
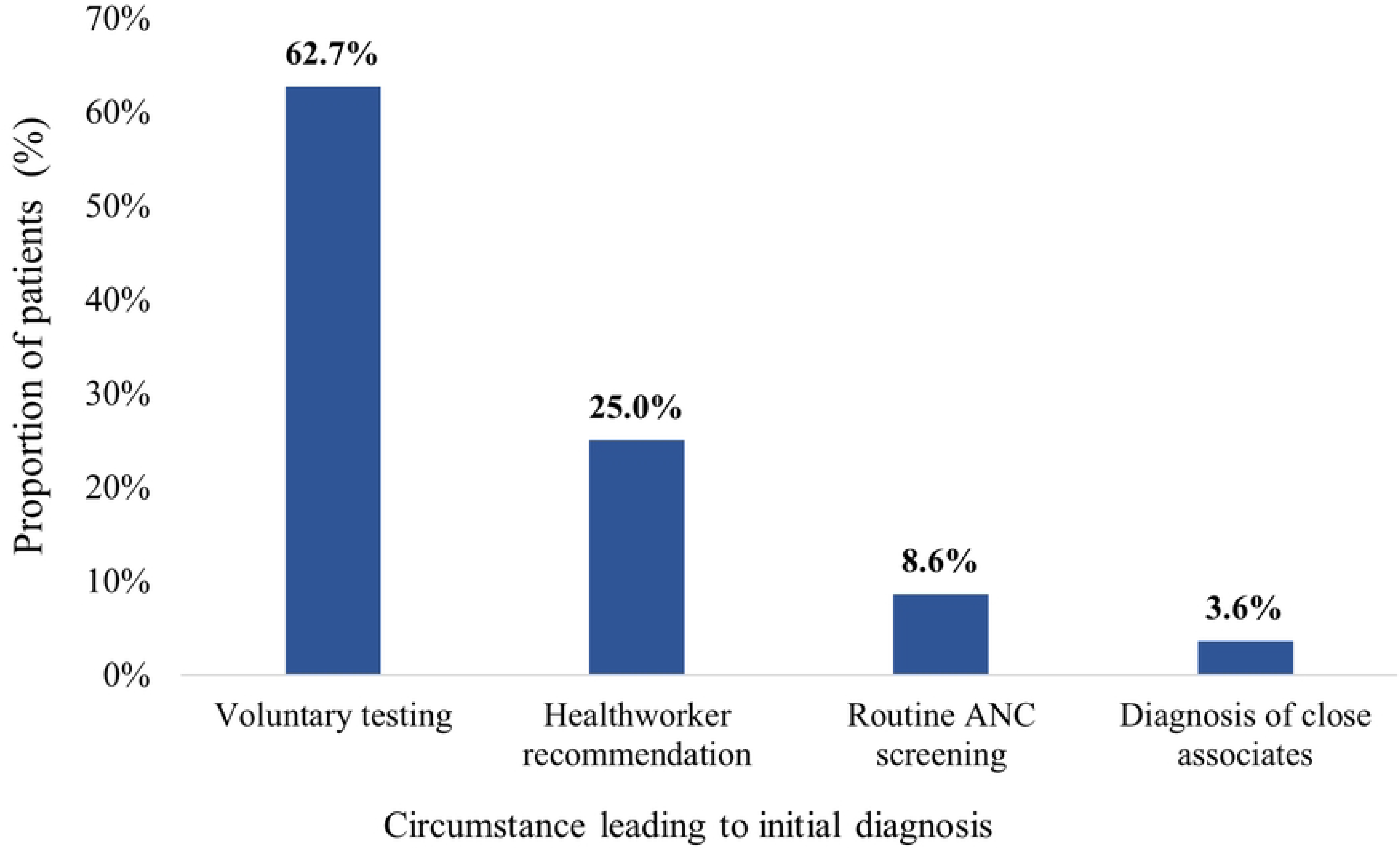
Circumstances leading to initial diagnosis among study participants (n = 220) ANC: antenatal care.

Approximately 60.9% did not know the HBsAg status of their immediate family members whereas 20.5% had a positive family history of HBV infection. In all, 15 (6.8%) had parents with HBV infection as illustrated in Fig 3.

**Fig 3.**
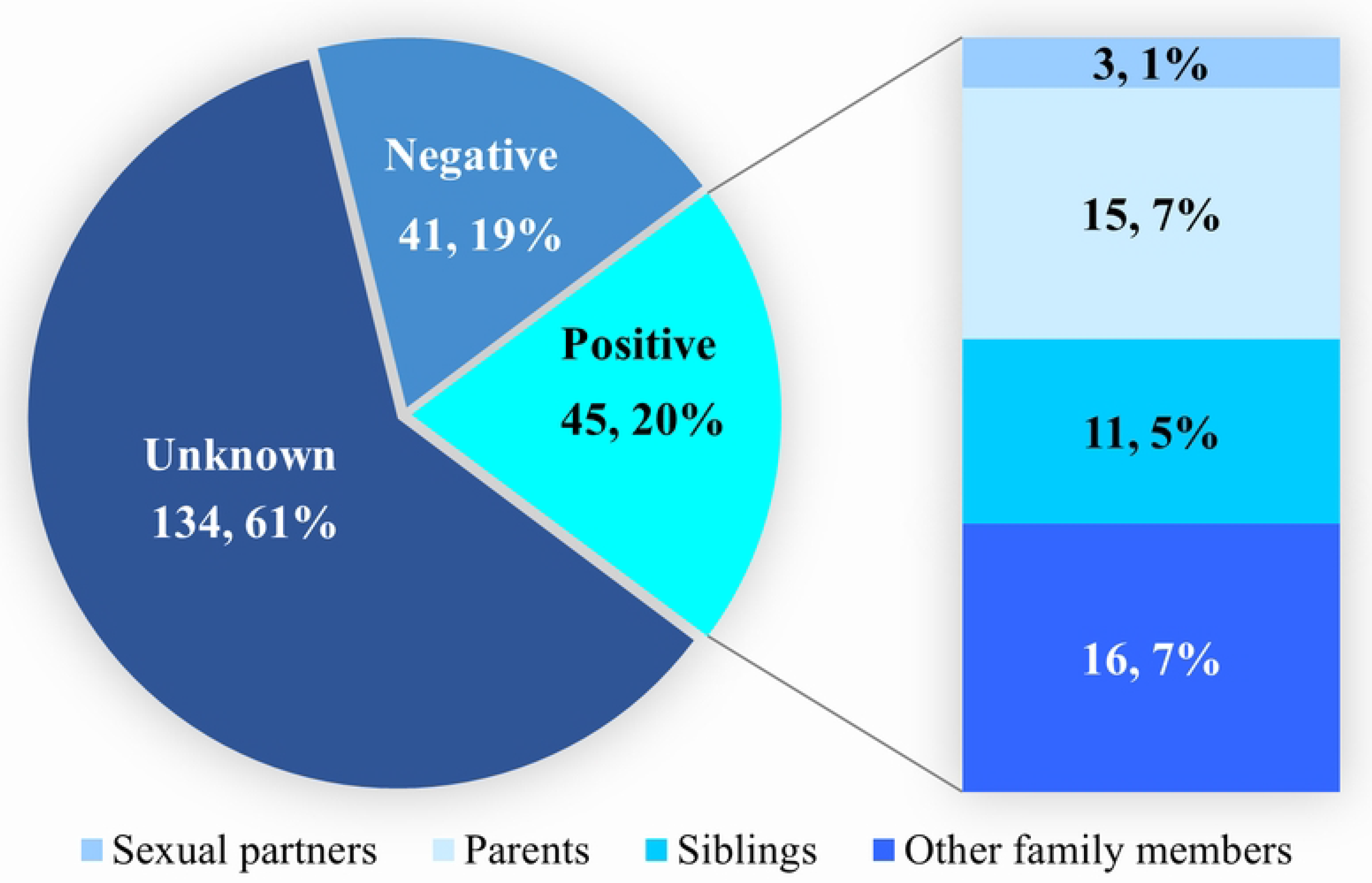
Distribution of HBV seropositive family members and sexual partners of the study participants.

In all, 24 patients (10.9%) had abnormal clinical examination findings. There were 12 patients (5.5%) who had icterus (jaundice), whereas 3 (1.4%) had hepatomegaly as illustrated in Fig 4 which shows the distribution of clinical examination findings among patients. Two patients (0.9%) each had abdominal distension and other stigmata of chronic liver disease (CLD).

**Fig 4.**
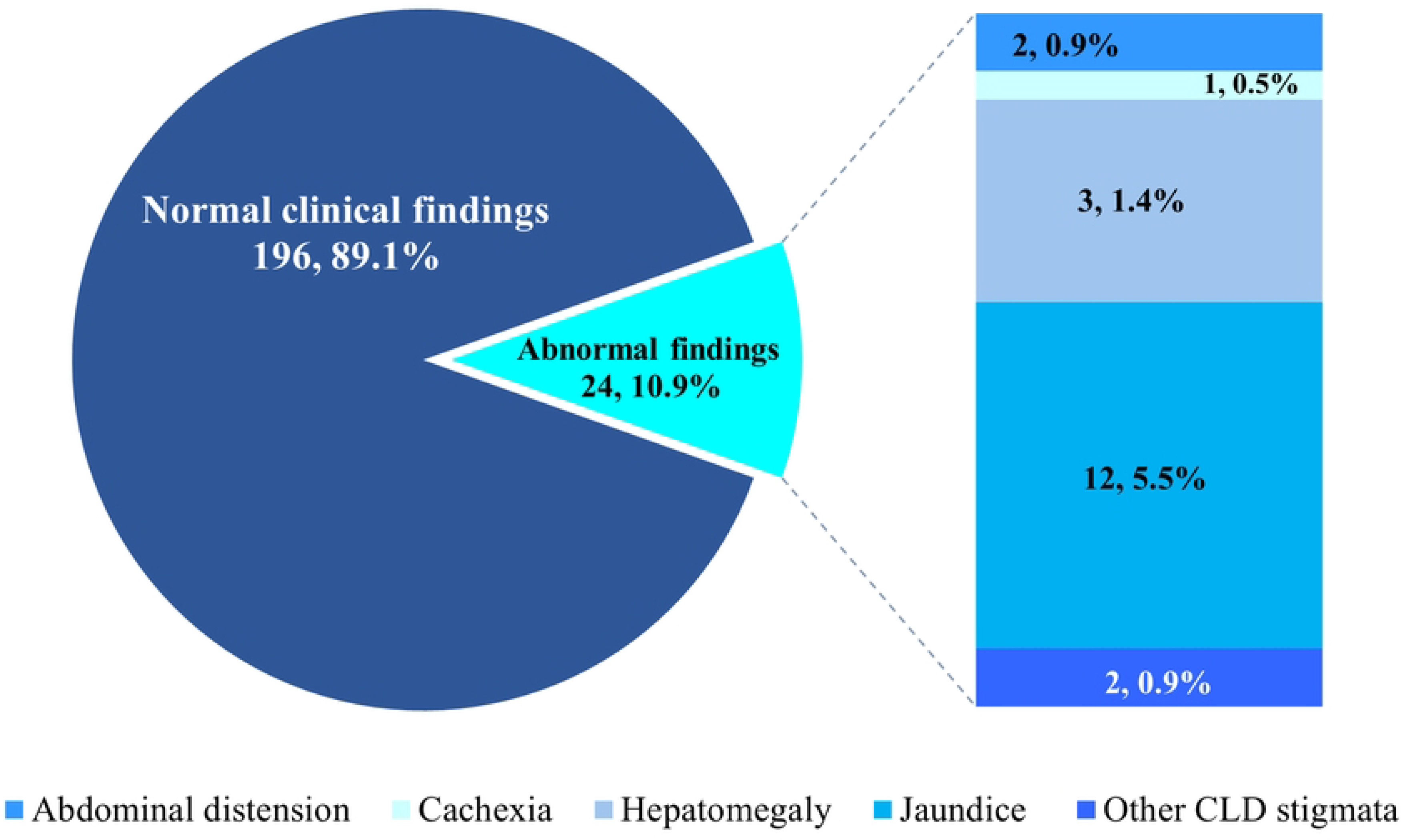
Distribution of clinical examination findings among patients. CLD = chronic liver disease.

### Laboratory characteristics of patients

Most of the participants had aspartate transferase (AST) and alanine transaminase (ALT) levels within the normal range, 131(59.6%) and 195 (88.6%) respectively. One hundred and eighty participants (81.8%) also had alkaline phosphatase (ALP) levels within the normal range as shown in Table 2.

**Table 2.**
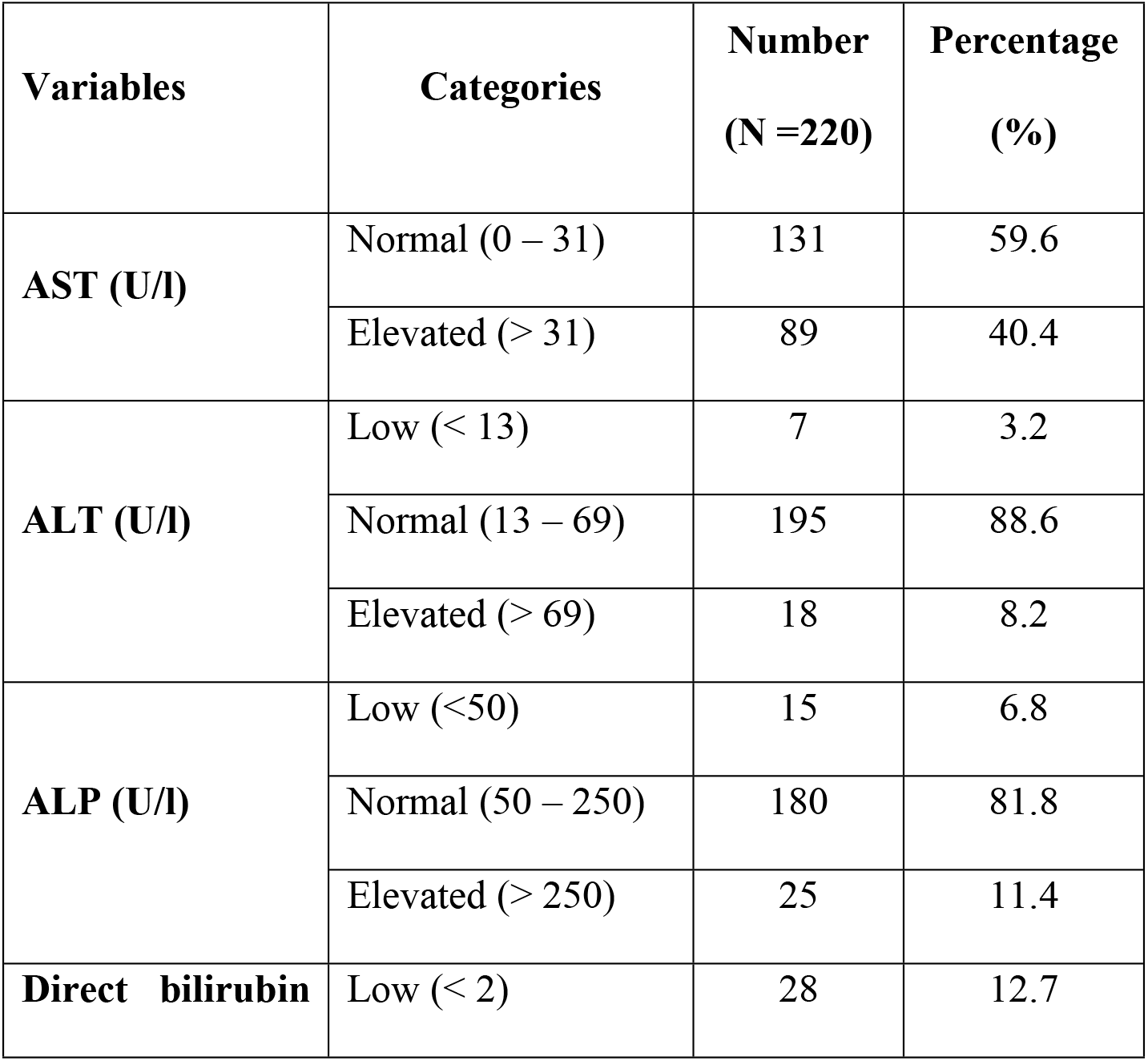
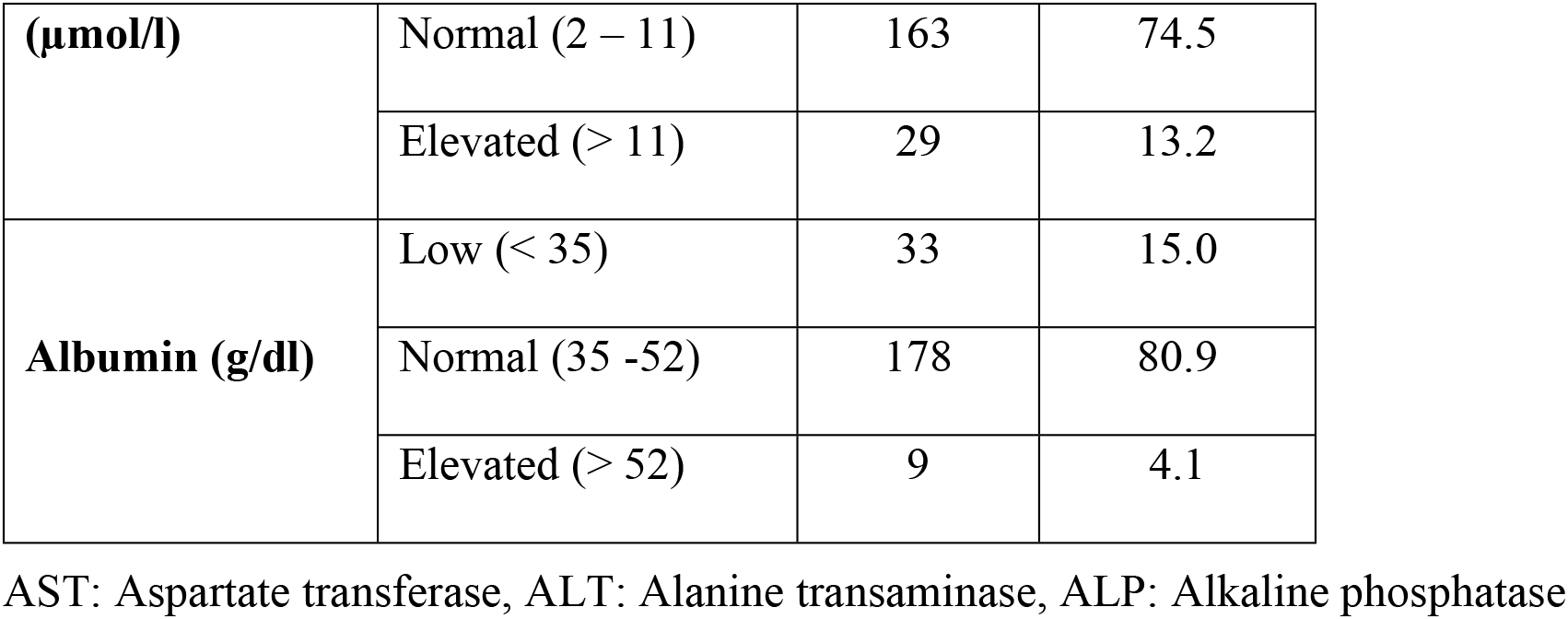
Liver function tests of study participants.

There was a small proportion of patients who had concomitant HIV and HCV infections, 2.7% and 1.4% respectively. Most of the patients (n = 85, 187%) had aspartate - platelet ratio index (APRI) scores > 0.5. Also, 172 patients (78.2%) had HBV DNA levels ≤ 2000 IU/ml whereas 21.8% (n = 48) had levels > 2000 IU/ml. Only one patient (0.5%) had an HBV DNA titer > 20,000 IU/ml (Table 3).

**Table 3.** Other laboratory parameters of study participants.

| Clinical parameters | Variables | Number | Percentage (%) |
| --- | --- | --- | --- |
| **HIV Status** | Positive | 6 | 2.7 |
|  | Negative | 214 | 97.3 |
| **HCV Status** | Positive | 3 | 1.4 |
|  | Negative | 217 | 98.6 |
| **APRI score** | < 0.5 | 33 | 15 |
|  | 0.5 – 2.0 | 186 | 84.5 |
|  | > 2.0 | 1 | 0.5 |
| **HBV DNA (IU/ml)** | < 2000 | 172 | 78.2 |
|  | 2000 – 20,000 | 47 | 21.3 |
|  | > 20,000 | 1 | 0.5 |

HIV: human immunodeficiency virus, HCV: hepatitis C virus, APRI: aspartate-platelet ratio index, HBV: hepatitis B virus, DNA: deoxyribonucleic acid

The proportion of patients who tested positive for HBsAb and HBcAb were 6 (2.7%) and 170 (77.3%) respectively. Also, 8 (3.6%) were seropositive for HBeAg whereas 177 (80.5%) showed seropositivity for HBeAb (Fig 5).

**Fig 5.**
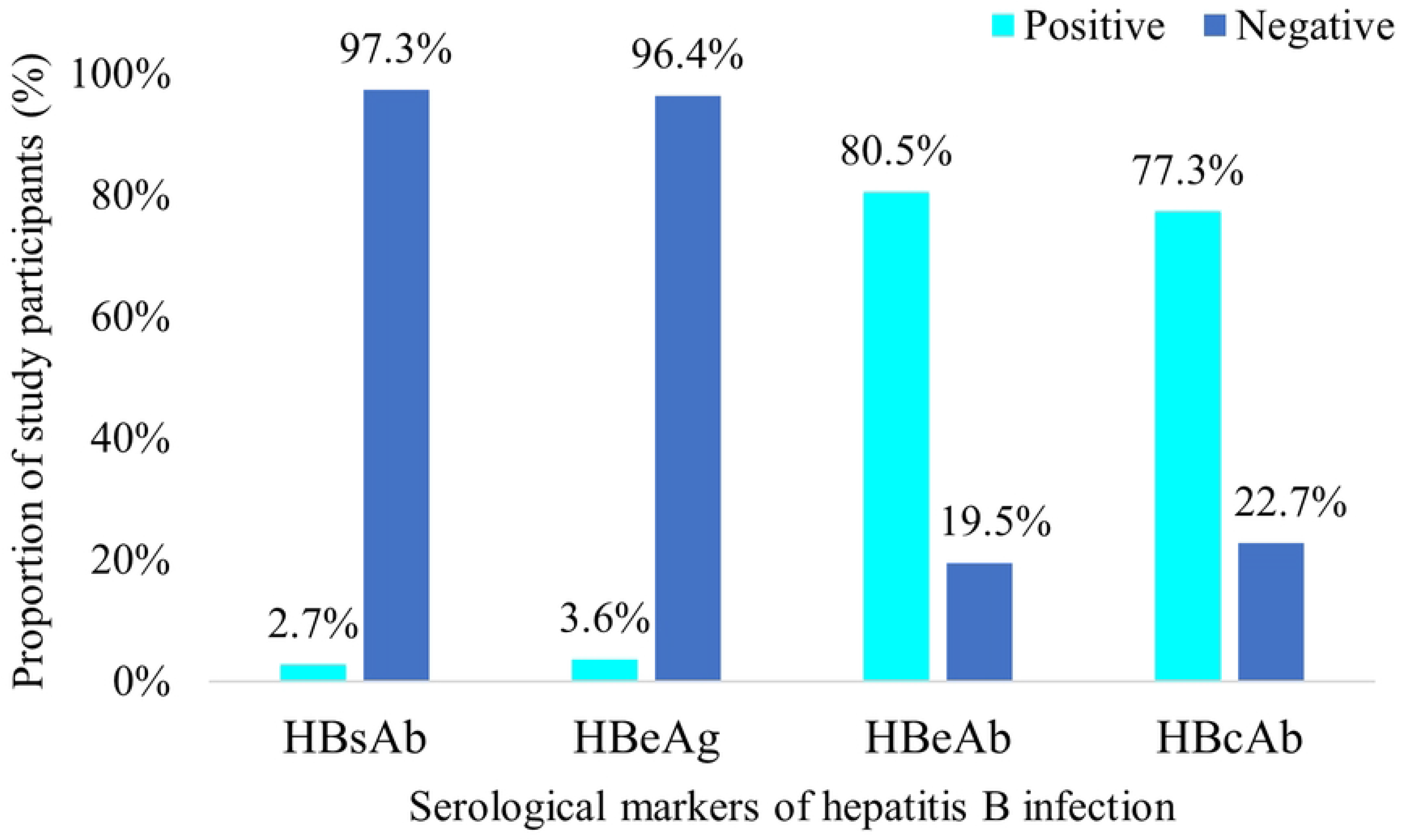
HBV serological profile of patients. HBsAb: hepatitis B surface antibody, HBeAg: hepatitis B envelope antigen, hepatitis B envelope antibody, hepatitis B core antibody

### Treatment Eligibility

In all, 14 participants (6.4%) were eligible for immediate initiation of antiviral therapy based on WHO guidelines [23]. This group comprised 8 (57.1%) males and 6 (42.9%) females (Table 4). All patients with e-antigen positive chronic hepatitis and/or APRI > 2 (HBV related cirrhosis or advanced fibrosis) required treatment as well as individuals with HCV/HIV coinfection and HCC. Among HBeAg-positive patients, treatment was required if ALT > 2x ULN or viral load ≥20,000IU/ml. For HBeAg-negative patients, indications for treatment were ALT ≥ 2x ULN or viral load ≥2000IU/ml. None of the pregnant females in the study were found to be eligible for immediate commencement of antiviral therapy for HBV infection.

**Table 4.** Characteristics of patients who were eligible for antiviral therapy.

| Antiviral treatment eligibility | Number n/N | Percentage (%) |
| --- | --- | --- |
| No antiviral treatment required | 206/220 | 93.6 |
| Treatment required: | 14/220 | 6.4 |
| Sex |  |  |
| Male | 8/14 | 57.1 |
| Female | 6/14 | 42.9 |
| HBeAg status |  |  |
| Positive | 8/14 | 57.1 |
| Negative | 6/14 | 42.9 |
| Serum ALT (U/l) |  |  |
| $\leq 2$x ULN | 0/6 | 0 |
| $> 2$x ULN | 6/6 | 100 |
| Pregnant women (n/N = 6/14) |  |  |
| Requiring treatment or prophylaxis | 0/6 | 0 |
| Not requiring treatment or prophylaxis | 6/6 | 100 |
| Advanced liver disease |  |  |
| APRI > 2 | 1/14 | 7.1 |
| Cirrhosis on USG | 0 | 0 |
| HCC on USG | 0 | 0 |
| No cirrhosis or HCC | 13 | 92.9 |
| Co-infection status |  |  |
| HIV-HBV co-infection | 6/14 | 42.9 |
| HCV-HBV co-infection | 3/14 | 21.4 |

HBeAg: hepatitis B envelope antigen, ALT: Alanine transaminase, ULN: upper limit of normal range, APRI: aspartate-platelet ratio index, USG: ultrasonography, HCC: hepatocellular carcinoma, HIV: human immunodeficiency virus, HBV: hepatitis B virus, HCV: hepatitis C virus.

## Discussion

This study aimed to describe the clinical, immunological, and virological characteristics of newly diagnosed HBV clients and assess their eligibility for antiviral treatment based on the current WHO guidelines [23]. The median age of the participants in this study of 34 years is comparable to the results of previous studies that were also focused on adult HBV patients ≥ 18 years. In a meta-analysis of 30 peer reviewed publications with a population pool of over 100,000 Ghanaians, the authors reported the highest prevalence of HBV infection amongst patients aged between 16 and 39 years [13].In sub-Saharan Africa, HCC has been reported to develop at a mean age of 33 years [24].Individuals aged between 31 and 40 years have been linked with a five-fold increase in the risk of HBV infection relative to other age groups (P=0.009). The age range reported in this study represents both the reproductive and most productive age groups in the Ghanaian society and may imply loss of productivity due to absence from work for hospital visits and other economic activities among these patients.

There were 42 more females than males in our study population, representing 19.1% of the total number of study participants. Two previous studies also reported a trend of higher numbers of females compared to males in their studies involving patients with hepatitis B infection [25, 26]. This study found a female to male ratio of 1.5:1 which can be partly accounted for by the disparity in the general health seeking behavior among both sexes [27]. Women are more at risk of contracting STIs than men [28]. This translates into a higher likelihood of disease transmission from men to women than vice versa [28]. Another study confirmed the existence of gender disparity in the prevalence of infections caused by HBV [29]. HBV infection is classified as a viral sexually transmitted disease together with other infections such as HIV, HCV, HPV and HSV. Majority of the patients (65.0%) were sexually active whereas the remainder were inactive. Sexual contact is key in the transmission HBV infection among adults in areas with very high prevalence rates including Ghana. In Ghana, mother-to-child transmission, unprotected intercourse, and transfusion of contaminated blood are the three main ways that HBV is spread [30].

In our study, we found that 63.3% of the patients did not have any knowledge about the status of their partners. This finding is not in keeping with the current WHO recommendation of routine screening of partners of all patients diagnosed with HBV [31]. Infection of an individual with HBV puts all close associates and immediate contacts such as spouses, sexual partners, and household members at a high risk of getting infected. Disclosure of one’s positive HBsAg status to such exposed close contacts is vital in the prevention or alleviation of the risk of transmission of HBV infection.

The disclosure of positive HBV status is very important for several reasons. First, it provides an opportunity for the diagnosed individual to receive social support [32]. Close contacts may be more likely to observe social protocols that will reduce their risk of being infected (such as not sharing personal items like razor blades, nail cutters, tooth brushes or anything that could have trace amounts of blood on them) and might be even more open to HBV vaccination. Further studies are needed to explore the impact of HBV status disclosure on the adoption of preventive measures and vaccination uptake among family members and close associates of infected patients in the Ghanaian setting. Knowledge of the positive diagnosis of a close contact is also an indication for HBsAg testing which will help in detecting positive cases in family groups. Disclosure can also motivate other seropositive family members to seek care thereby reducing their chance of developing cirrhosis and HCC.

In a study published in 2020 that investigated the disclosure of CHB status among HBV infected patients in Ghana, it was reported that patients were more likely to disclose their status under some conditions and for some reasons than others [33]. Reasons for disclosure included the desire to have close contacts tested and vaccinated, willingness to confide in the target of disclosure and the need for social and/or financial support. On the contrary, non-disclosures were on account of phobia for stigmatization and past undesirable experiences associated with disclosure.

Most of the participants were initially diagnosed via voluntary testing (62.7%) followed by health worker recommendation (25.0%). Similar findings were reported in a previous study that demonstrated that individuals with HBV infection were diagnosed because of self or physician-initiated testing, testing during screening programs and ANC testing [26, 34]. The findings of this study indicate that voluntary testing for HBsAg among the adult population is very important in HBV infection case detection. Community based or opportunistic screening undoubtedly remains an effective tool in picking up positive cases in the general population.

In our study, we found that majority of the patients (60.9%) were not aware of the HBsAg status of their immediate family members. Out of those who knew the status of their family members (n=85), majority (n=45, 52.9%) had family members who were known to be seropositive for HBsAg. Therefore, the actual number of patients with a positive family history for HBV infection is likely to be far greater. These findings suggest that intrafamilial spread of HBV infection may be a potential mode of transmission in Ghana, and further studies are needed to fully elucidate this theory.

The overall prevalence rate of HBsAg positivity was 20.5% among household members of patients which is comparable to the rate of 23.3% reported in Iran [35]. A 2024 study in Mwanza, Tanzania however reported a much lower HBV prevalence rate of 5.4% among household contacts of index cases [36]. After testing positive for sexually transmitted diseases like HIV and HBV, some individuals are more prone to concealing their diagnosis than seeking medical attention due to stigmatization [37]. CHB infection is an incurable disease and hence, some people may not want to be associated or tagged with this diagnosis. A qualitative study on chronic hepatitis B stigma in Ghana found that people with hepatitis B experience societal stigma underlined by the belief that HBV is a highly contagious and severe disease as well as the notion that HBV infection arises due to curses [38]. The question of “the most probable source of transmission” arises when individuals test positive for STIs. Some HBV positive patients may not wish to disclose their status to their close contacts which will prompt them to get tested. If this testing turns out positive, they would likely be viewed as the source of the infection in the household simply because they were diagnosed first, though it could be the other way around. Nondisclosure of HBsAg positive status to close contacts increases their risk of contracting the disease. It also robs them of an indication for testing and treatment as well as possibly HBV vaccination if eligible.

The median time interval between patient’s initial diagnosis and presentation at the specialist clinic was 8.5 months (IQR 3.0-22.5). Majority (61.8%) were seen < 1 year whereas 8.6% were seen > 5 years after initial diagnosis. The reasons for this delay in presentation at the specialist clinic may be associated with both patient and primary care provider factors. In several African countries including Ghana, many patients who test positive for HBsAg during opportunistic screening are unable to afford the additional tests or the cost of therapy because it must be paid for out-of-pocket [10, 13, 39]. As a result, a lot of patients who test positive for HBV do not proceed immediately to seek medical care or do not complete the required initial evaluation to determine what needs to be done for them. In Ghana, anecdotal evidence suggests that some patients resort to traditional and/or herbal remedies for the initial management of STIs such as HIV, HCV, and HBV as well as other health conditions including cancer, diabetes, hypertensive heart disease and bronchial asthma[40-42]. As a result of this exploration and experimentation with unorthodox medicine it is possible that some patients may present late to the hospital after all other avenues have failed or when there is disease progression.

According to the WHO, as of 2019, 10.5% of all the people estimated to be HBsAg positive already knew their positive status whereas only 22% of those diagnosed were receiving medical care [4]. This points to the fact that a huge proportion of HBV infected patients do not receive medical care after their diagnosis. Additionally, certain impediments to HBV treatment and care have been reported including barriers relating to sociocultural beliefs and health systems. These sociocultural beliefs include the perception that HBV is a spiritual disease and a form of punishment from God. On the individual level, some patients take HBV infection for granted owing to the absence of specific symptoms of CHB [33]. Furthermore, the financial burden of hospital-centered treatment for HBV infection is considerably high and even prohibitive for some patients [43]. In Ghana, unlike patients with HIV or HBV/HIV coinfection, HBV mono-infected patients must pay for their medications out of pocket in addition to the cost of their routine investigations such as LFT, HBV immunological profile and hepatitis B viral load. The inclusion of medications, laboratory analysis and other investigations under the National Health Insurance Scheme (NHIS) could potentially remove this barrier at least partially if not completely. Similar to findings in other countries such as Burkina Faso and Australia, the delay in seeking care for HBV infection could also be because of stigmatization (or fear of it thereof), lack of knowledge and misconceptions about HBV treatment as well as long queues and waiting times at treatment facilities [44, 45].

Hepatitis B infection is known to be self-limiting in 95% of adult cases and 5-15% of cases among children under five years. According to WHO, 6.6 million (22%) of the people diagnosed with HBV were on treatment as of 2019. The percentage of patients diagnosed with HBV in Ghana who require treatment as well as the proportion of these treatment eligible patients who receive the required treatment is unknown. In 2021 the proportion of CHB patients worldwide who required treatment was about 12 - 25% (WHO 2021). There is no specific treatment for acute hepatitis B unless the patient has acute hepatic failure, but for most patients, antiviral treatment for CHB is lifelong. This has direct cost implications and potential financial toxicity.

Treatment for viral hepatitis in Ghana is based on the National Guidelines for Prevention, Care and Treatment of viral hepatitis which is in line with the treatment recommendations of the World Health Organization. Approved antiviral agents for treatment of HBV infection in Ghana include Tenofovir disoproxil fumarate, Entecavir, pegylated interferon and Emtricitabine among several others. The treatment eligibility rate of 6.4% reported in this study is much lower than the reported rate of 46.8% in the United States [46]. This low rate implies that a significant proportion of the patients were diagnosed at a stage of the disease that required close monitoring but did not warrant the immediate initiation of expensive antiviral therapy which is not affordable for many patients.

Early and appropriate initiation of therapy for HBV infection is important in the reduction of the risk of transmission and development of fatal long-term complications. To be able to accomplish the WHO goal of eliminating viral hepatitis by the year 2030, it is important to not only precisely estimate treatment eligibility rates but also to ensure high treatment rates. To the best of our knowledge based on currently available literature this is the first study that evaluates treatment eligibility (rate) among HBV infected patients seeking medical care in the Central Region of Ghana. Evaluation of treatment rate i.e., proportion of patients eligible for treatment who receive antiviral treatment was beyond the scope of this study and remains an outstanding question for future studies.

### Limitations

This study was conducted among patients who had voluntarily reported at the HBV/other STIs clinic to seek medical care for HBV infection and thus did not include in-patients admitted at the medical emergency unit or on the medical wards for complications of hepatitis B infection. Neither did the study capture HBV patients who were not seeking hospital-based care at all. Thus, the results of this study may potentially underestimate the prevalence of advanced disease among HBsAg seropositive patients.

## Conclusions

This study reports a high level of HBsAg positivity rate among close family members of HBV positive persons. Nonetheless, there is very low adherence to the requirement of screening of close family members of index cases. There is a very low treatment eligibility rate among patients seeking medical care for HBV infection. There was low prevalence of both HIV and HCV coinfections without any recorded cases of triple infections.

## Data Availability

All relevant data are within the manuscript and its Supporting Information files.

